# Factors Influencing Mental Health During Covid-19 Outbreak: An Exploratory Survey Among Indian Population

**DOI:** 10.1101/2020.05.03.20081380

**Authors:** Absar Ahmad, Ishrat Rahman, Maitri Agarwal

## Abstract

**Purpose:** Research on the impact of social distancing on mental health during epidemics is limited, especially in India. The purpose of this study is to scale the association between anxiety and socio-demographic factors during Covid19 lockdown among the general Indian population.

**Design/methodology/approach:** A descriptive cross-sectional nationwide study was designed to enrol the general population. The inclusion criteria for this study were Indian citizens aged 18 years and above. The study was conducted from 29^th^ March to 12^th^ April 2020, using an online google questionnaire. The anxiety among respondents was detected and measured using a Generalised Anxiety Disorder Scale which consists of 7 questions (in English), i.e. GAD-7.

**Findings:** Respondees were 392, and from these participants, the prevalence of anxiety was 25.3 per cent. Based on the bivariate logistic regression analysis, the predictors of anxiety were gender, religion, occupation as business/self-employed, marital status, family size, health status and sleep deprivation.

**Conclusion:** This study reports the prevalence of anxiety among Indian population who were grounded at their homes during lockdown due to coronavirus pandemic in the country.

**Limitations:** (1) The selection of participants through non-random sampling. (2) Because of the cross-sectional character of the study, causal conclusions cannot be drawn.

**Originality/Value:** This paper fulfils an identified need to study the mental health status of the population under situations like lockdown, thereby helping fill a persistent gap in Indian research on this issue.

## Background

As the increase in stringent measures to keep people apart through lockdown to slow the spread of the coronavirus disease 2019 (COVID-19) pandemic (1) comes, a profound, disturbing effect on all aspects of society, including mental health and physical health can be anticipated (2). Anxiety is the most common mental health disorders in the general population and can be characterised by the feelings of tension and worrying thoughts (3). Evidence suggests that people with anxiety disorders are at higher risk of developing a number of chronic medical conditions (4). The prevalence of anxiety differs from social and cultural factors as well as in different classes of ethnic groups (5).

The prevalence of anxiety has been reported previously among isolated people due to Middle East Respiratory Syndrome(6). The core symptoms of anxiety include excessive and uncontrollable worries, sleep disturbances, and difficulty in concentrating (7). During disease outbreaks, community anxiety can rise following the first death, increased media reporting, and an escalating number of new cases (8). A study evaluated the psychological outcomes of people who were quarantined, which is an extreme form of social distancing, during outbreaks of SARS, H1N1 flu, Ebola and other infectious diseases since the early 2000s. They found that individuals experienced both short term and long term mental health problems (9). For instance, one particular study compared quarantined versus non-quarantined individuals during an equine influenza outbreak, reporting high levels of anxiety and depression (34%), during the outbreak compared with only 12 per cent in non-quarantined individuals (10).

India is the 2^nd^ most populated country in the world, and currently, more than 1.3 billion people are in lockdown since 25^th^ March. The government took this step in fear for an outbreak in the country since it is densely populated, the result could have been catastrophic (11), if not taken timely and followed strictly. However, these measures are the largest of their kind in the world. They risk heaping further hardship on the quarter of the population who live below the poverty line and the 1.8 million who are homeless (12).

As a whole, India is facing lockdown for the first time in its history. To date, there is no profound evidence showing how quarantine affects anxiety among the general population during isolation in India. However, there are a few reports from Kashmir which are facing complete lockdown since August 2019, describing an increase in patients experiencing anxiety, stress sufferings, loneliness, frustration and abnormalities in behaviour (13). So there is an urgent need for studies to investigate the impact of lockdown during outbreaks, like coronavirus on mental health (14). This study is a small step taken to analyse the effect of lockdown among the Indian population, and it was envisioned to highlight the importance of research in the area of isolation and mental health, which is among the less touched issues in India. Thus, the current study was designed to provide a valuable addition to the epidemiology of mental stress among the general population across India.

The objectives of the study were to (i) estimate the prevalence and (ii) explore sociodemographic and health-related correlates of anxiety disorders among Indians during Covid19 lockdown.

## Material and Methods

**Design:** This study adopted a descriptive, cross-sectional questionnaire-based survey. During the time of lockdown, the only method for data collection possible was online. Thus the present study used an online survey that includes quantitative research methods. This online survey has the advantage of using the Internet to provide researching from a distance (15); additionally, it saved time and cost (16).

**Sampling Technique:** A non-random convenience sampling method was used. Participants were selected from the contact list of the first researcher, being invited by WhatsApp and email to complete the online survey. The online survey lasted for two weeks, from 29^th^ March to 12^th^ April 2020. The inclusion criteria were citizen of India and age greater than 18 years. The consent form was also included in the online survey tool regarding their participation in the study. The distribution of the questionnaire was extended to the whole country, however mostly in Uttar Pradesh to where the first researcher belongs. The response was also received from Indians staying abroad at present and facing lockdown.

**Sample Size:** The prevalence of anxiety disorders was 3.6 percent according to National Mental Health Survey (17) in the general population in India while the prevalence of anxiety according to the Global Burden of Study was 3.3 percent (18). However, these studies were not conducted at the time of an emergency, when anxiety among people is considered to be at a peak. Thus to calculate sample size, we selected the maximum sample size, which comes at p=0.5. Subsequently, the sample size was determined by using the formula Z^2^pq/d^2^ (‘p’ is the prevalence of anxiety which is taken as 0.5, ‘Z’ is the type 1 error at 5 per cent, and ‘d’ is the absolute error). Therefore, based on sample size calculations (p=0.5, q=1-p, Z=1.96, d=0.05), the requisite random and representative sample size was 384. However, surveys were collected from 392 participants.

## Development of Study Tool

The current research was conducted after reviewing the literature available on the mental health of a population in isolation. The Google Forms are used for designing and developing web-based questionnaires that are automatically hosted via a unique URL. This URL link gave people round the clock access from anywhere in the world. The responses were secured using “Cloud” database where the data was automatically sorted, scaled and scored by custom Excel formulae. The researcher could download real-time questionnaire responses in multiple formats (e.g. excel) which were then analysed with a statistical software of choice. The free availability of the tool and automatic recording of user responses in its spreadsheet had made data collection and analysis simple. In a country like India, where the internet user base is increasing day by day web-based survey tools became an obvious choice for survey research (19,20).

**Exposure variables:** The survey questionnaire included socio-demographic variables such as age, gender, place, education, occupation, religion, family income, marital status and number of family members. Any change in the amount of sleep and use of mobile during lockdown was also recorded. Health-related variables such as self-reported health status with three-point scale and response options were ‘poor’, ‘average’ and ‘good’. Since the literature suggests anxiety is more extensive in the low religiosity subgroup than in the high or no religiosity subgroup (21), questions related to religiosity were asked, with a four-point scale with options as ‘not at all’, ‘somewhat’, ‘very’ and ‘extremely’. These questions were ‘How much do you participate at a religious ceremony’, ‘How much do you turn to the higher presence (e.g. Allah, God)’ and ‘How much do you read religious /Spiritual Books’(22).

## Outcome variables

Generalised anxiety disorder (GAD) is one of the most common mental disorder, and it often remains undetected (23). Thus, several screening instruments have been developed to measure anxiety. One of these instruments is the Generalised Anxiety Disorder Scale GAD-7. The 7-item Generalized Anxiety Disorders Scale (GAD-7) was developed as a screener for generalised anxiety disorder (GAD) in primary health care setting (24). Psychometric evaluations of the GAD-7 suggest that it is a reliable and valid measure of GAD symptoms in the general population (23,25) as well as individuals isolated due to risk of infection (6). The GAD-7 has demonstrated good psychometric properties, including sensitivity and specificity for diagnosing GAD (24). The present study has used the English version of the GAD-7, and internal consistency was assessed by using Cronbach’s α. The internal reliability of the present study was found to be 0.87. For each of 7 items, subjects were asked about how frequently they felt each one during the lockdown period. The 4-point *Likert scoring system* was used as follows: not at all (0 points), several days (1 point), more than half the days (2-point), nearly every day (3-point). Higher scores implied more significant anxiety symptoms. We use a GAD cut-off score of >10 for performing the binary logistic regression analyses to identify predictors of high anxiety (6).

## Data analysis

The data recorded in the spreadsheet were exported to SPSS. Descriptive and inferential statistics were used for the analysis. The odds ratios and their 95 per cent CI were calculated. A p≤0.05 was considered to be statistically significant.

## Ethical Consideration

Permission to conduct study was obtained from Institutional Ethics Committee of the Career Institute of Medical Sciences and Hospital, Lucknow. Personal identification like Name, contact number, email id was not asked due respect of participant’s privacy. Participation in this survey was completely voluntary and participants can withdraw at any time prior to the completion of the survey.

## Results

A total of 392 out of 407 participants completed the whole items in the GAD-7 questionnaire. Nine were excluded from the data analysis due to incomplete data and six were due to not fulfil the inclusion criteria. The study included 392 Indians from 24 states in the country and 10 from abroad as well. The age of the participants ranged from 18 to 71 years, with a mean age of 30.3(SD 9.28) years. About 61 % were below 30 years of age, and 47 % were female. More than 90 % were graduated, and about 64 % followed Hinduism as their religious belief, 30 % were Muslims, and 6 % included other religions. About 42 % of respondents were students, followed by private job holders (25 %), Government job holders (17.6 %) and Business/self-employed (8 %). About 59 % were never married, and 54 % of the respondent’s family size was 4 or less. Nineteen percent of respondents self-reported health status were poor and scored 6 or less in religiosity score (Table 2). About 15 % of the respondents reported a lack of sleep during the lockdown, and 60 % reported increased use of mobile.

**Table 1:** Participants geographical location.

| **Location** | **n** | **%** |
| --- | --- | --- |
| Central | 17 | 4.3 |
| East | 31 | 7.9 |
| North-East | 8 | 2.0 |
| North | 235 | 59.9 |
| South | 13 | 3.3 |
| West | 78 | 19.9 |
| Outside India | 10 | 2.6 |
| **Total** | **392** | **100.0** |

*Central*-Madhya Pradesh, Chhattisgarh; *East*-Bihar, Jharkhand, Orissa, West Bengal; *North East*-Assam, Meghalaya, Manipur, Sikkim; *North*- Jammu and Kashmir, Delhi, Haryana, Punjab, Uttar Pradesh, Uttarakhand ; *South*-Karnataka, Kerala, Tamil Nadu ; *West*-Gujarat, Goa, Maharashtra, Rajasthan ;*Outside India*-Saudi Arabia, Hungary, Thailand, Brazil, Oman, Italy

**Table 2:** General characteristics of the respondents (n=392)

|  |  | **n** | **%** |
| --- | --- | --- | --- |
| Age | ≤ 30 | 237 | 60.5 |
|  | > 30 | 155 | 39.5 |
|  | mean±SD | 30.3±9.28 |  |
| Gender | Female | 185 | 47.2 |
|  | Male | 207 | 52.8 |
| Education | Intermediate and less | 32 | 8.2 |
|  | Graduate | 168 | 42.9 |
|  | PG and More | 192 | 49.0 |
| Religion | Hindu | 252 | 64.3 |
|  | Muslim | 118 | 30.1 |
|  | Other | 22 | 5.6 |
| Occupation | Students | 162 | 41.3 |
|  | Private salaried | 99 | 25.3 |
|  | Government salaried | 70 | 17.9 |
|  | Business/self-employed | 31 | 7.9 |
|  | Other | 30 | 7.7 |
| Monthly family Income(in INR) | ≤20,000 | 52 | 13.3 |
|  | 20,000-50,000 | 94 | 24.0 |
|  | 50,000-1,00,000 | 102 | 26.0 |
|  | 1,00,000-5,00,000 | 91 | 23.2 |
|  | >5,00,000 | 53 | 13.5 |
| Marital Status | Never Married | 232 | 59.2 |
|  | Ever Married | 160 | 40.8 |
| Family size | ≤4 | 212 | 54.1 |
|  | >4 | 180 | 45.9 |
|  | mean±SD | 4.72±2.1 |  |
| Self-Reported health status | Poor | 76 | 19.4 |
|  | Average | 179 | 45.7 |
|  | Good | 137 | 34.9 |
| Religiosity | ≤6 | 226 | 57.7 |
|  | >6 | 166 | 42.3 |
|  | mean±SD | 6.2±2.0 |  |
| Lack of Sleep | No | 333 | 84.9 |
|  | Yes | 59 | 15.1 |
| Uses of Mobile increases | No | 155 | 39.5 |
|  | Yes | 237 | 60.5 |
| **Total** |  | **392** | **100.0** |

*Source: Online primary survey*

Table 3 presents the prevalence of anxiety with background characteristics. The prevalence of anxiety among respondents was 25.3 %, based on the cut-off point of 10 and above on the GAD- 7 scale. A total of 99 participants were found facing anxiety in this study. Occupation, monthly income, marital status, family size, self-reported health status, and sleep were found associated with anxiety. However, age, gender education, religiosity and use of mobile were not found to be associated with anxiety in the bivariate analysis.

**Table 3:** Prevalence of anxiety with background characteristics.

|  |  | **Prevalence**<br>**n,(%)** | **$\chi^2$, P value** |
| --- | --- | --- | --- |
| Age | ≤ 30 | 63(26.6) | 0.55, 0.455 |
|  | > 30 | 36(23.2) |  |
| Gender | Female | 53(28.6) | 2.13,0.144 |
|  | Male | 46(22.2) |  |
| Education | Intermediate and less | 9(28.1) | 2.28,0.320 |
|  | Graduate | 48(28.6) |  |
|  | PG and More | 42(21.9) |  |
| Religion | Hindu | 48(19.0) | 16.9,0.000 |
|  | Muslim | 46(39.0) |  |
|  | Other | 5(22.7) |  |
| Occupation | Students | 44(27.2) | 12.15,0.016 |
|  | Private salaried | 20(20.2) |  |
|  | Government salaried | 15(21.4) |  |
|  | Business/self-employed | 15(48.4) |  |
|  | Other | 5(16.7) |  |
| Monthly family Income(in INR) | ≤20,000 | 15(28.8) | 10.45,0.033 |
|  | 20,000-50,000 | 27(28.7) |  |
|  | 50,000-1,00,000 | 21(20.6) |  |
|  | 1,00,000-5,00,000 | 30(33.0) |  |
|  | >5,00,000 | 6(11.3) |  |
| Marital Status | Never Married | 68(29.3) | 4.95,0.026 |
|  | Ever Married | 31(19.4) |  |
| Family size | ≤4 | 40(18.9) | 9.97,0.002 |
|  | >4 | 32.8(59) |  |
| Self-Reported health status | Poor | 31(40.8) | 13.42,0.001 |
|  | Average | 43(24.0) |  |
|  | Good | 25(18.2) |  |
| Religiosity | ≤6 | 54(23.9) | 0.524,0.469 |
|  | >6 | 45(27.1) |  |
| Sleep decreased | No | 77(23.1) | 5.32,0.021 |
|  | Yes | 22(37.3) |  |
| Increased uses of Mobile | No | 38(24.5) | 0.074,0.785 |
|  | Yes | 61(25.7) |  |
| **Total** |  | **99(25.3)** |  |

Source: Online primary survey

Table 4 displays the predictors of anxiety. After adjusting for other factors in the bivariate logistic regression model, the male gender was negatively associated with anxiety (OR=0.50, 95% CI:0.287–0.883, p<0.05). Muslim participants had 2.48 times higher risk of developing anxiety as compared to Hindu participants (OR=2.48, 95% CI:1.371–4.517, p<0.01). The results also showed that participants who reported occupations as Business/self-employed were 3.75 times higher risk of developing anxiety, as compared to those who reported their occupation as student. The odds of developing anxiety were almost 40 % lower among ever married participants compared to never-married participants. Participants with average health status (OR=0.457, 95 % CI:0.239–0.873, p<0.01) and good health status (OR=0.402, 95 % CI:0.190–0.847, p<0.01) have a lower risk of developing anxiety as compared those who reported poor health status. Loss of sleep was significantly associated with anxiety. Participants who reported a loss of sleep had 1.97 times higher risk of anxiety (OR = 1.98, 95 % CI: 1.012–3.889, p < 0.05).

**Table 4:**
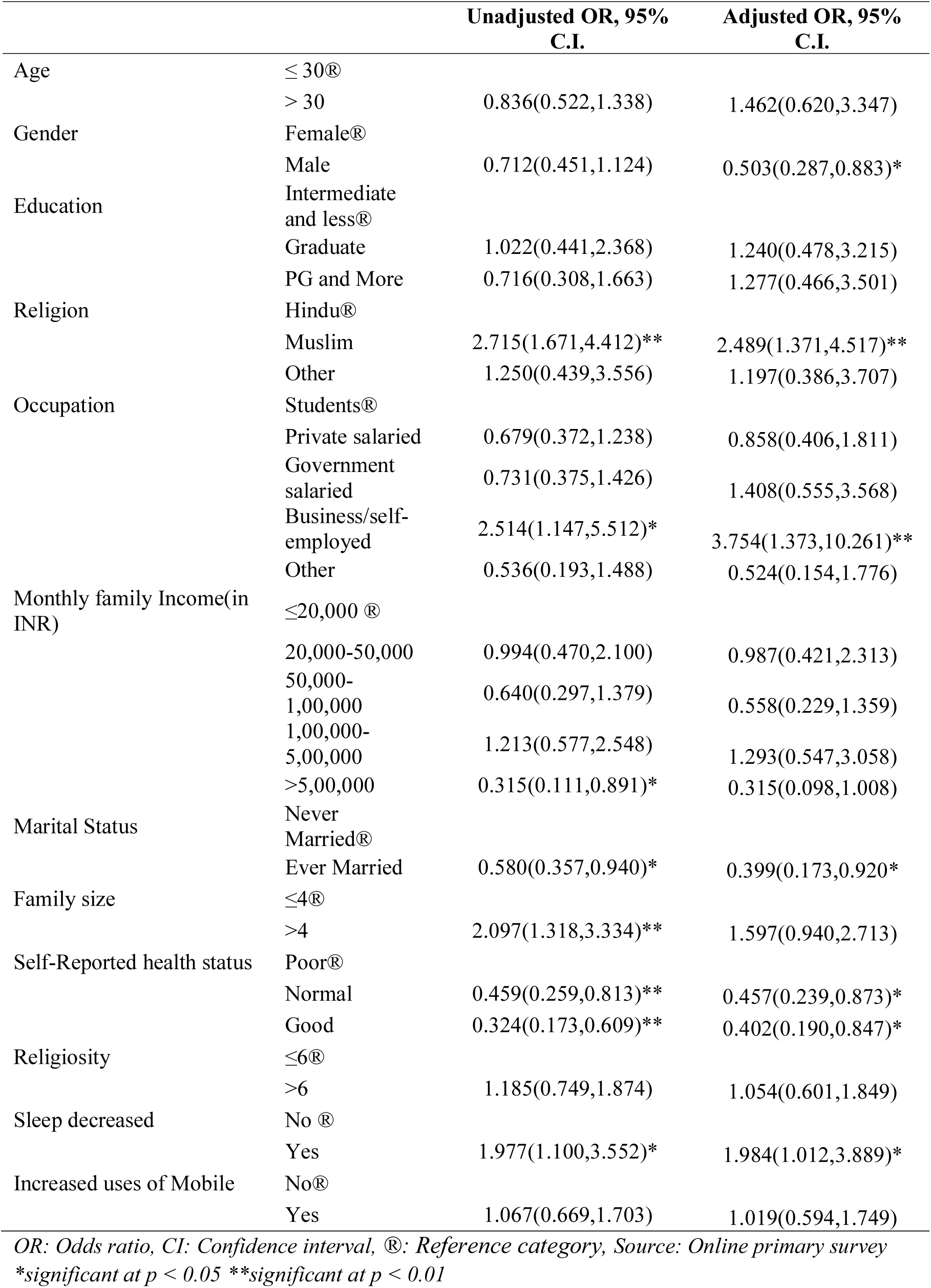
Bivariate logistic regression analyses for predicting anxiety.

## Discussion

People stressed due to longer quarantine duration as well as infection fears, frustration, boredom, inadequate supplies, inadequate information, financial loss, stigma (9) as well as anxiety, which may turn into depression and high perceived stress (3).

The results in this study indicate that 25.3 % of the participants are facing anxiety, which is very high from prevalence (3.6%) estimated in National Mental Health Survey (2015–16) in the general population (17). While in the study of the mental health status of isolated people due to Middle East Respiratory Syndrome (MERS) in South Korea found the prevalence of anxiety using GAD-7 to be 7.6 %, which is one-third of the prevalence in this study (6). In another study of psychological distress determined by the Kessler 10 Psychological Distress Scale among a population affected by highly infectious Equine influenza in Australia, the anxiety was reported to be 34% (10). Uncertainty captures the ideas of fear for one’s health or the health of loved ones, disruption to daily life, and the unknown aspect of disease (26). The reason behind the high prevalence of anxiety among Indians could be because it is the first encounter of this type of lockdown. Besides, this pandemic is impacting a population already facing challenges in their lives like unemployment, family issues and various other changes like lifestyle, which is included as a significant reason for anxiety (14).

The current study determines a lower rate of anxiety among males. Other studies in India (27), (28) as well as in Palestine (29), Germany (23), UK (30) and US (31),, also report males to have a lower degree of anxiety. Globally, the prevalence of anxiety disorders was almost double in females (5.2 %) as compared to males (2.8 %) estimated using 272.2 million people in 2010 (32). One explanation of why women tend to be more prone to stress is because they ruminate about life stressors, which can increase their anxiety, while men engage more in active, problem-focused coping (33). In India, it is common for females to serve the family, and especially during the lockdown, females are managing household chores and office work at home. They have to keep up with the demands of all the family members constantly, such as food and cleaning, as well as home-schooling children, who are also not able to get their regular education due to the lockdown. Thus, rendering females physically and mentally exhausted, which may contribute to higher anxiety levels. Other reasons may be that women are more likely to experience physical and mental abuse than men (33), and abuses of any kind are found to be linked to the development of anxiety disorders (33).

Occupations such as Business/ Self-employed are also found to be predictors of anxiety in the present study. Currently, industries like tourism, textile, and agriculture along with the employment generated through this are at greater risk (34). In total, about 100 million and more Indians’ jobs are at risk during and even beyond the Covid-19 lockdown. Retailers of non-food items have closed their outlets, and food retailers are also expecting loss (35). One of the risk factors of anxiety among isolated people in South Korea due to MERS was financial loss (6). Fear of financial loss was higher among businessmen or self-employed individuals.

In the present study, Muslims were found to be at a higher risk of anxiety compared with Hindus. One reason for higher anxiety among Muslims was repeated hate crimes in recent years (36). Furthermore, the health ministry repeatedly blamed an Islamic seminary group gathering for spreading the coronavirus, and governing party officials spoke of “human bombs” and “corona jihad” resulting in a spree of anti-Muslim attacks across the country (37). There is evidence from study indicated a strong positive relationship between sociocultural adversities and psychological distress(38). Another reason could be, about half of the Indian Muslims are self-employed (39), so they are at more risk of financial loss compared to their counterparts. Although more research is needed to confirm these results with different samples of Muslims, the present study sheds important light on a topic that has not been previously examined.

In earlier studies, marital status was not shown to influence levels of Anxiety (40), but our study showed that unmarried people have more anxiety than ever-married people. Similar findings were also registered in Western Australia, New South Wales (41) and US (42). Economic down-turn affects adult unemployment as well as education, and this could be a contributing factor as to why unmarried people experience a higher level of anxiety. The total participants were made up of 42 % students, and most of them were unmarried. Students are likely to be anxious and worried about their education especially their exams, and their career since the economy has taken a downturn.

This study evaluated the low presence of GAD among those who reported their health status as either average or good. The thought of being in poor health (43) likely makes them more prone to anxiety. This lends credence to prior studies that suggest close associations between health status and anxiety (42). Lower levels of self-reported health status may give rise to sleep disturbances, fatigue, pain that may trigger worry and anxiety. The inverse association between anxiety and health status also raises the possibility that anxiety may cause poorer health status. We postulate that the presence of anxiety may be associated with lower compliance with medical treatments, thus undermining health (42). Also, anxiety among our sample may be because patients are facing problems in accessing health services amid lockdown (44).

Our study reported higher anxiety issues in those who reported sleep deprivation. Similar findings were also reported in the German Population using GAD-7 (23). The sleep pattern of people altered due to self-isolation and routine changes. A change in the amount of time spent outside could most likely influence the routine at home, for instance, time of awakening and sleeping and time of a meal. Many people may feel more fatigue related to the mental workload associated with covid-19, and this fatigue can also be caused by psychological states, such as stress and anxiety (45). The pandemic has made people confused and uncertain and given some a sense of trepidation. All these feelings can lead to poor sleep quality, which in turn can make people more tired and anxious (42).

Our study has some limitations, including sampling strategy, which is non-random sampling limiting the generalizability. Another limitation was that the causality relationship could not be entirely ascertained because the design of the study is cross-sectional. Further studies with a larger sample size may shed more light on this issue. Other limitations pertain to the study’s dependent measure, i.e. anxiety. The cut-point used in the present study may have misclassified a certain percentage of individuals.

Despite the limitations mentioned above; this study is an important step in the field of this type of research and has policy implications. The findings suggest a need for more research to better understand the epidemiology of anxiety among people during emergence. In addition to epidemiologic investigations on prevalence and risk factors of anxiety, concerted efforts are needed to be put forth into developing evidence-based healthy coping techniques and problem-solving skills for people at risk.

## Conclusion

The present study was able to determine the psychological impact of Covid-19 lockdown on the Indian population. This study was made with the sight to collect mental health-related data among the Indian population during lockdown amid disease outbreak, and it may have been done for the first time in the current scenario. Study findings indicate that the Indian population under lockdown is affected by heightened psychological distress and thus higher prevalence of anxiety. Statistical analysis indicated that certain groups were more vulnerable to anxiety; specifically, female, Muslims, self-employed, never married, reported poor health and reported poor sleep.

## Data Availability

All data underlying the results are available as part of the article and no additional source data are required.

